# Pulsed Field Versus Radiofrequency Ablation For Repeat Ablation of Atrial Fibrillation: Evaluation of Outcomes

**DOI:** 10.64898/2026.10.08.26365096

**Authors:** Vishal Patel, Carmel Ashur, Joseph Adewumi, Kishan Padalia, Matthew M. Zipse, Alexis Z. Tumolo, Lohit Garg, Syed Rafay Ali Sabzwari, Amneet Sandhu, Michael A. Rosenberg, Wendy S. Tzou

## Abstract

**Background:** Pulsed-field ablation (PFA) has recently been approved in the USA as a new method for treating atrial fibrillation (AF). PFA provides a promising new approach in the treatment of atrial fibrillation (AF), which historically has a high recurrence rate post-ablation. Of significance, PFA has shown promising data in terms of safety and procedural efficiency to radiofrequency ablation (RFA).

**Objective:** The purpose of this study was to compare the procedural and health outcomes in patients with recurrent AF undergoing ablation using PFA or RFA.

**Methods:** Consecutive patients with a history of previous radiofrequency ablation and recurrent AF undergoing redo ablation with PFA or RFA were included. Patients were seen for follow-up in the clinic ranging from 4 months to a year and we reported ablation strategy and arrhythmia type. The primary outcome was recurrence of any atrial arrhythmia after a 90-day blanking period.

**Results:** A total of 242 patients with recurrent AF underwent redo ablation using PFA (n = 54, 22.5%) or RFA (n = 188, 77.5%). The PFA cohort was younger on average (66 vs. 70 years) with a lower mean CHA2DS2-VASc score (2.2 vs. 3.06, p=0.001) compared to the RFA cohort. PFA procedures were shorter (162±53 vs 206±60 minutes, p<0.001) but required more fluoroscopy (13.9±8.9 vs 8.7±6.3 minutes, p<0.001). Among patients seen on follow-up, recurrence occurred in 28% of PFA versus 55% of RFA patients, with longer arrhythmia-free survival after PFA (log-rank p=0.03). PFA remained associated with lower recurrence after multivariable adjustment (HR 0.4, 95% CI 0.2–0.9, p=0.04). Adverse events, predominantly vascular access–related, were similar.

**Conclusion:** Among patients with recurrent atrial arrhythmias after prior RFA, repeat PFA was associated with shorter procedures, greater fluoroscopy use, and lower arrhythmia recurrence, with comparable safety.

## Introduction

Pulsed-field ablation (PFA) has transformed the global practice of catheter ablation for atrial fibrillation (AF), especially since the first systems received approval from the United States Food and Drug Administration in late 2023.^1–3^ Clinical trials have demonstrated PFA to be non-inferior in treating persistent or paroxysmal AF, compared to thermal energy sources.^3^ Additional studies have suggested that PFA may have a more favorable safety profile,^4–7^ with respect to risk of esophageal or phrenic nerve injury or pulmonary vein (PV) stenosis,^5^ and greater efficiency in achieving pulmonary vein isolation (PVI), leading to rapid world-wide adoption and virtual supplanting use of traditional point-by-point radiofrequency ablation (RFA), especially for first-time AF ablation.^1,2,5,8,9^ Additionally, while initial PFA catheter designs were tailored for PVI, use has also extended to target non-PV sources for AF, such as the posterior wall (PW) and superior vena cava,^10–14^ also with relative efficacy and safety compared to thermal energy sources. Although freedom from atrial arrhythmias (AAs) has been similar to those achieved with RFA or cryoablation, significantly reduced residual AA burden has been more often observed among patients undergoing PFA versus RFA or cryoablation.^15^

Repeat AF ablations have been reported among 8.8% of patients previously treated with PFA, versus 15-30% of those in whom thermal energies were utilized.^16–18^ However, although initial limited remapping post-PFA studies suggested durable PVI in as many as 91-100% at 3-6 months, registry data reflecting real-world practice have demonstrated PV reconnections in 55-57%. Results regarding durable PW isolation following PFA have been similar, with PW reconnections reported in 54-55%.^17,18^ As use of PFA for AF ablations continues to increase, questions regarding efficacy and safety continue to be raised.^5,19,20^

Limited data exist comparing the efficacy of PFA versus RFA in for repeat AF.^21^ We sought to investigate outcomes at our center among patients who had previously undergone RFA for AF treatment and who underwent repeat AF ablation using RFA versus PFA.

## Methods

### Patient population

The study population consisted of consecutive patients ≥ 18 years old with history of prior RFA for paroxysmal or persistent AF and who underwent repeat ablation for recurrent atrial arrhythmias using either RFA or PFA, at the University of Colorado Anschutz Medical Campus January 2022-November 2024. PFA was not available for clinical use at the University of Colorado until May 2024. The patients were divided into 2 groups based on the ablation energy used during repeat CA. Data collection and analysis were approved by the institutional review board of the University of Colorado School of Medicine.

### Electrophysiology Mapping and Ablation Procedures

All procedures were performed with patients in fasting state and under general anesthesia. Antiarrhythmic drugs were held for at least 5 half-lives for most procedures, with the exception of amiodarone, which was held variably at operator discretion. Oral anticoagulation was continued without interruption. Ultrasound-guided common femoral venous access was obtained. Diagnostic catheters included a decapolar catheter positioned in the coronary sinus for reference (6F Inquiry, Abbott, Plymouth, MN; 6F WEBSTER™ Decapolar Catheter, Johnson & Johnson MedTech, Irvine, CA; or Dynamic XT™, Boston Scientific, Marlborough, MA), a multipolar mapping catheter (PENTARAY™, OCTARAY™, or OPTRELL™, Johnson & Johnson MedTech, Irvine, CA; Advisor HD Grid or HD Grid X, Abbott, Plymouth, MN), which was used to map the right atrium (RA) and left atrium (LA), and a phased-array intracardiac echocardiography (ICE) catheter (8F Soundstar, Johnson & Johnson MedTech, Irvine, CA, or 9F ViewFlex, Abbott, Plymouth, MN). Heparin was administered as a bolus and infused to achieve and maintain activated clotting times >350 s prior to transseptal puncture and for the duration of LA access. Transseptal puncture (1-2, at operator discretion) was performed using RFA needle (NRG Transseptal Needle, Boston Scientific, Marlborough, MA) or RFA wire (VersaCross Connect, Boston Scientific, Marlborough, MA) via steerable sheath (8.5F CARTO Vizigo, Johnson & Johnson MedTech, Irvine, CA; 8.5F or 13F Agilis, Abbott, Plymouth, MN; or FARADRIVE, Boston Scientific, Marlborough, MA), and under ICE and fluoroscopy guidance. If 2 transseptal punctures were performed, the second transseptal sheath was a non-steerable sheath (8F SL1, Abbott, Plymouth, MN). Electroanatomic mapping was performing using CARTO (Johnson & Johnson MedTech, Irvine, CA) or Ensite Precision or Ensite X (Abbott, Plymouth, MN). Baseline atrial voltage and activation maps were acquired in presenting or atrial-paced rhythm unless the presenting rhythm was AF, in which case direct-current cardioversion was performed to achieve sinus rhythm prior to mapping. Patients presenting in atrial flutter (AFl) or atrial tachycardia (AT) underwent initial activation mapping in tachycardia.

Chronic reconnections from prior ablation were determined by voltage or activation mapping, to assess sites of entrance conduction, and pacing (output 10-25mA, pulse width 2ms), to assess sites of exit conduction. Reconnection sites were characterized by bipolar voltage >0.5 mV, earliest local activation time, and pace-capture within regions of prior ablation. In cases in which linear lesion sets had been previously delivered to treat LA or RA flutters, differential pacing was additionally utilized to determine sites of residual conduction. Ablation was then performed targeting sites of reconnection, with ablation energy chosen based on operator discretion.

Following ablation, programmed stimulation was performed following administration of isoproterenol (either administered in single bolus of 30 mcg or as an infusion of up to 30 mcg/min). Adenosine (3-24mg, with goal of achieving AV block or PR prolongation) was also infused, at operator discretion, to assess for dormant conduction following ablation. Additional mapping and ablation were performed if additional, sustained atrial arrhythmias were inducible or if acute reconnections or dormant conduction were observed.

### Radiofrequency Ablation Details

Esophageal temperature probes (Smiths Medical, Minneapolis, MN) were inserted at the outset of each RFA case. RFA was delivered using an open-irrigated, force-sensing catheter with 3.5-mm tip electrode (ThermoCool SmartTouch SF or QDOT Micro, Johnson & Johnson MedTech, Irvine, CA, or TactiCath or TactiFlex, Abbott, Plymouth, MN), using 40-50 W for 5-30s at non-PW locations or 40-90W for 4-7s at PW sites, with shorter durations implemented during higher power applications. Non-PW RFA lesions exceeding 7 seconds were guided by impedance and local electrogram (EGM) changes; on the PW, lesions at a given site were ceased for esophageal temperature rise of >0.1°C.

### Pulsed Field Ablation Details

All PFA procedures were performed with patients under general anesthesia and paralyzed and utilizing the FARADRIVE sheath or 13F Agilis (Abbott, Plymouth, MN),and 31-mm FARAWAVE PFA catheter (Boston Scientific). Catheter manipulations were guided over a 0.035-in Rosen wire (Cook Medical, Bloomington, IN). PV reconnections were targeted with the PFA catheter in basket or flower configuration, or both, at operator discretion, with 2 applications in each configuration for each site of PFA delivery. Non-PV reconnections, including linear ablation sites, were treated with the catheter in flower configuration, with 2 applications per site, and ensuring overlap of the territory of interest of the pentaspline electrodes. Atropine 0.4-1mg or glycopyrrolate 0.6mg were administered intravenously to prevent vagal reactions, and IV nitroglycerin 0.5-1mg was administered to prevent coronary vasospasm, when PFA was applied near the mitral or tricuspid valve.

### End points and follow-up

The primary outcome evaluated was atrial arrhythmia-free survival during follow-up. Any recurrence within a blanking period of 90 days was documented separately than recurrence following this period. Recurrence was defined as any episode of AF, typical or atypical atrial flutter, or atrial tachycardia with a duration of at least 30 seconds and persisting beyond the 90-day blanking period. Secondary efficacy endpoints included the total procedure duration and total fluoroscopy time. Patients were followed in outpatient clinic visits at 6-8 weeks, 3 months, 6 months, 12 months, and then annually following repeat ablation, or more frequently as clinically indicated. The visits included physical examination, 12-lead electrocardiogram (ECG), and ambulatory monitoring for at least 7 days using a patch monitor or continuously for those with an implantable loop recorder in place. For those with recurrence within the blanking period, data was collected up to 365 days post blanking period to evaluate future recurrence. This was then used to determine the correlation between recurrence within the blanking period and the following year after. Additional findings such as LA enlargement were determined based on echocardiogram readings obtained from patient EMR prior to procedure.

### Statistical analysis

Categorical variables are presented as numbers and percentages and compared using either the χ^2^ or Fisher’s exact test. A logistic regression was used to compare recurrence of AF in the setting of recurrence within the blanking period. Multivariate Cox PH analysis was used as a method of time-to-event analysis with consideration of multiple risk factors. This was used to isolate recurrence with specific ablation type with multiple variables. Hazard ratios were reported for individual factors influencing chance of recurrence with 95% confidence intervals (CI). The Kaplan-Meier analysis, together with the log-rank test, was used to compare the probability of freedom from atrial arrhythmia recurrence*. P* < .05 was considered statistically significant. The analysis was performed using R and RStudio.

## Results

### Demographics

A total of 242 patients with recurrent AF underwent redo ablation using PFA (n = 54, 22.3%) or RFA (n = 188, 77.7%). Patients receiving PFA were significantly younger (mean±SD age 66±12.3, versus 70±12.3 years), were less likely to have congestive heart failure (CHF) or hypertension and thus had lower CHADSVASC scores (2.2 ±1.6, vs. 3.1±1.6), although had larger left atrial (LA) sizes, compared to those undergoing repeat RFA (Table 1).

**Table 1:**
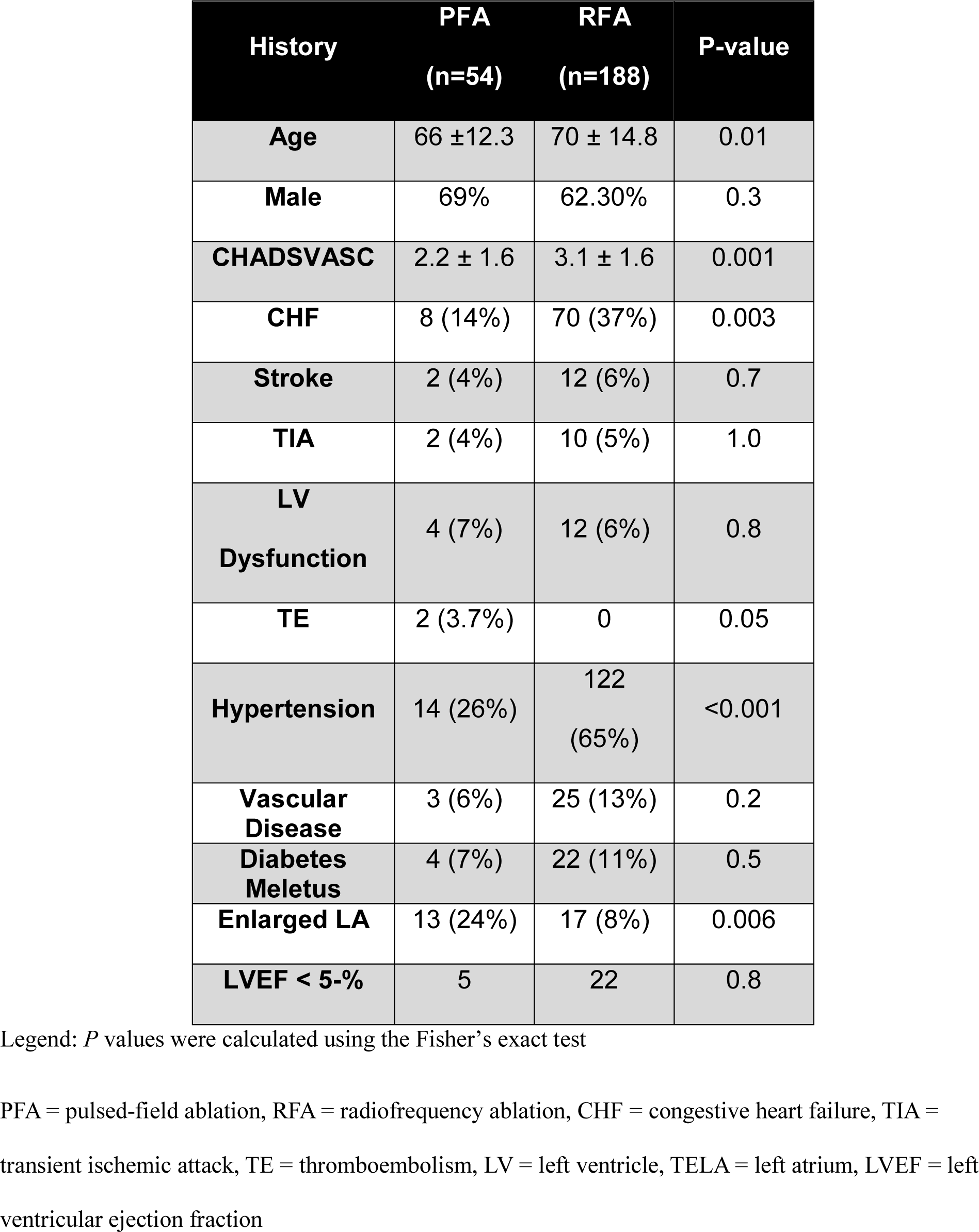
Demographics and baseline comparisons between PFA and RFA.

### Procedural Characteristics

Although fluoroscopy times were longer in PFA versus RFA procedures (13.9±8.9, versus 8.7±6.3 min, (p = 0.0002, Table 2 and Fig 1A), procedure times were shorter (161.9±52.5, vs. 205.9±60.4 min, p < 0.0001).

**Figure 1A:**
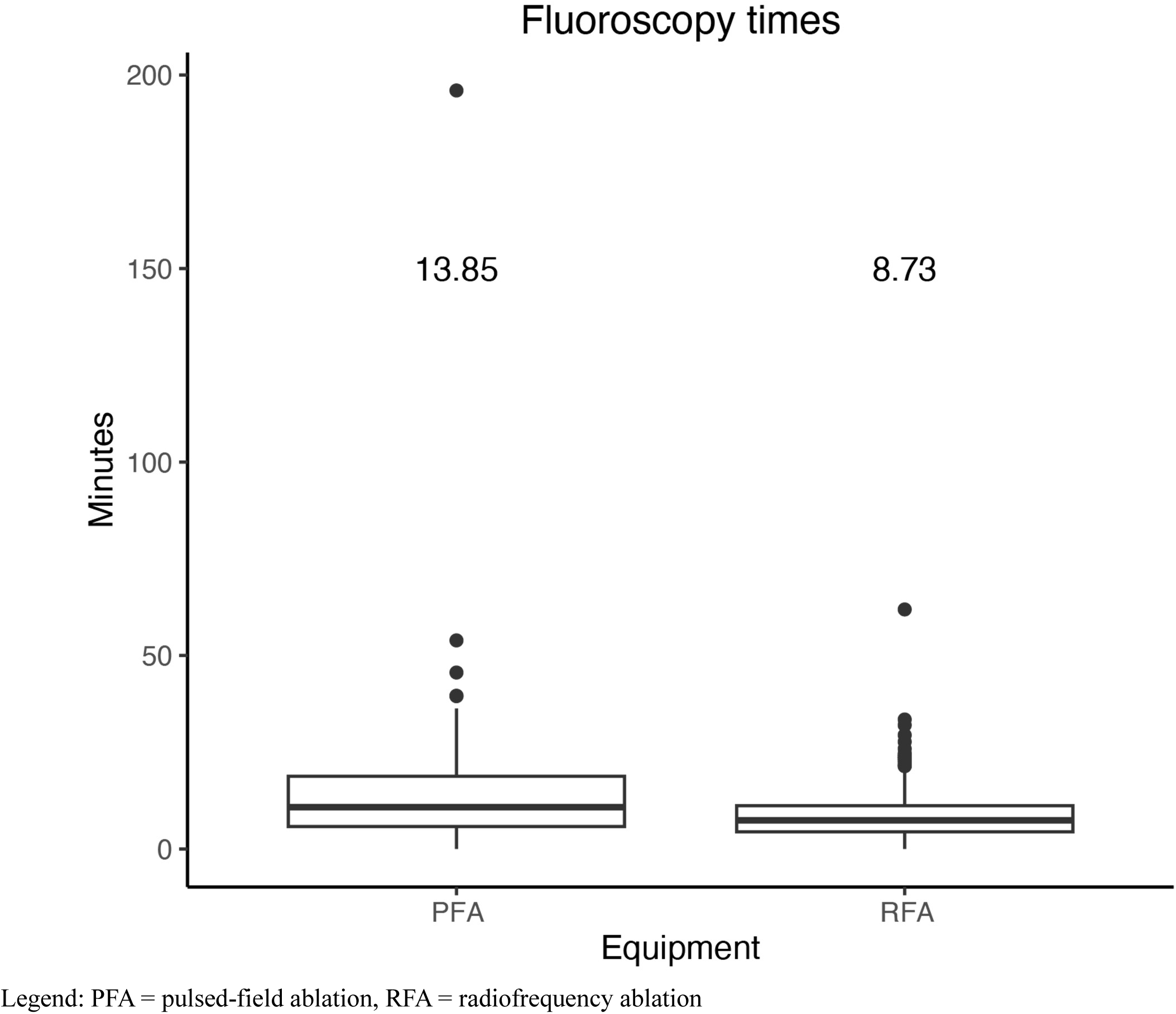
Fluoroscopy time. PFA = pulsed-field ablation, RFA = radiofrequency ablation

**Table 2:**
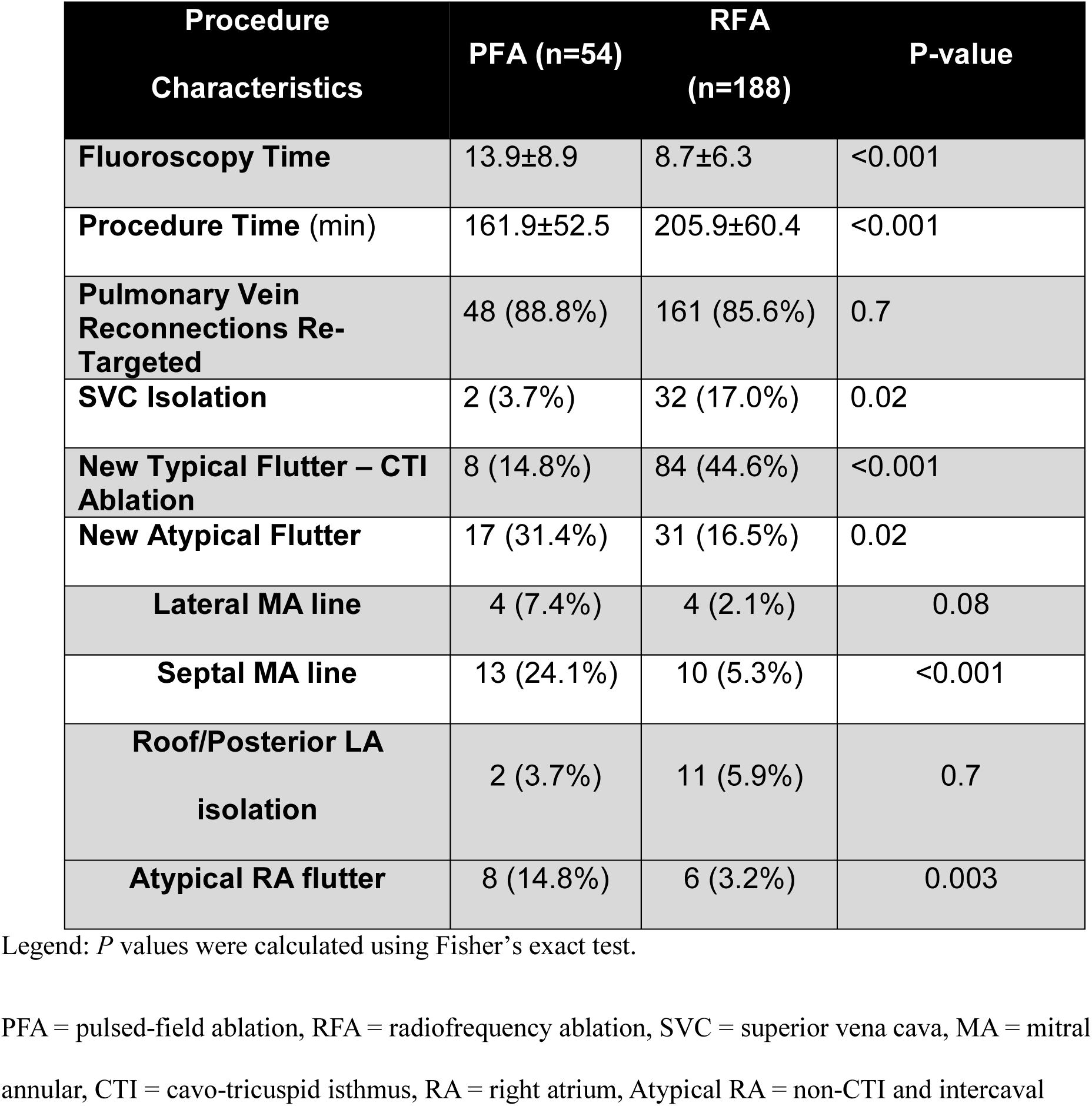
Procedure characteristics.

A majority of patients underwent additional non-PV ablation (Table 2, Fig 1B and 1C) to target non-PV AF triggers or atrial flutter (AFL). Superior vena cava (SVC) isolation was more often performed using RFA (17.0% vs. 3.7%, p=0.02). While the proportions undergoing non-PV ablation were similar between PFA and RFA groups, there were notable differences in AFL types targeted; typical AFL was more often targeted with RFA compared to PFA (44.7% vs. 14.8%, p<0.001), while the converse was true for atypical AFL (16.5% vs. 31.4%, p=0.02). Of note, mitral annular AFL was more likely to be targeted with anteroseptal mitral annular (MA) linear ablation overall (p = 0.01), but especially when PFA was used compared to RFA (Table 2).

**Figure 1B:**
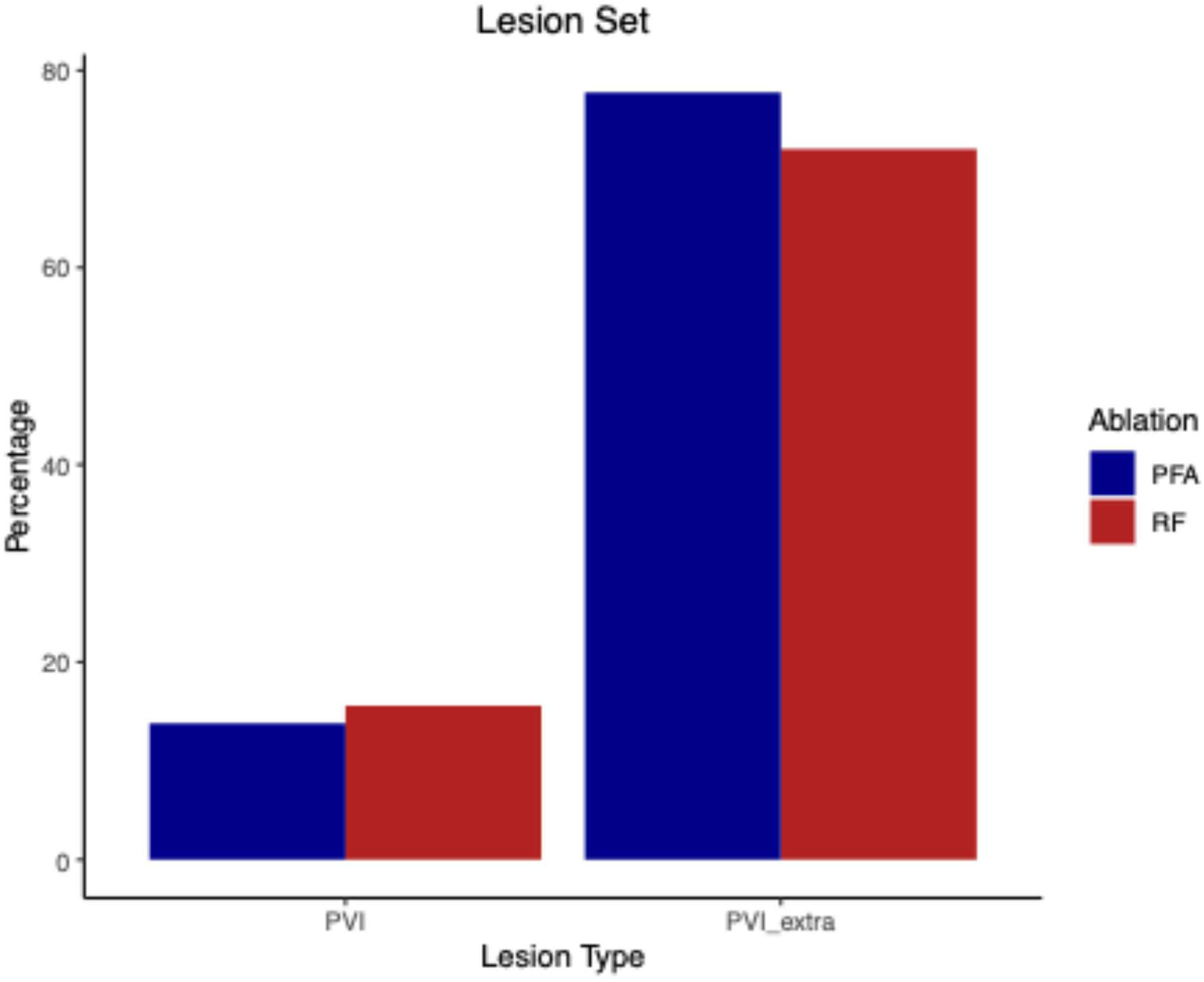
Lesion sets performed. PVI = pulmonary vein isolation only, PVI_extra = other lesions including cavo-triscupid isthmus (CTI), posterior wall (PW), mitral isthmus (MI), roof, mitral annulus (MA), right atrium (RA), intercaval IVC region, PFA = pulsed-field ablation, RFA = radiofrequency ablation

**Figure 1C:**
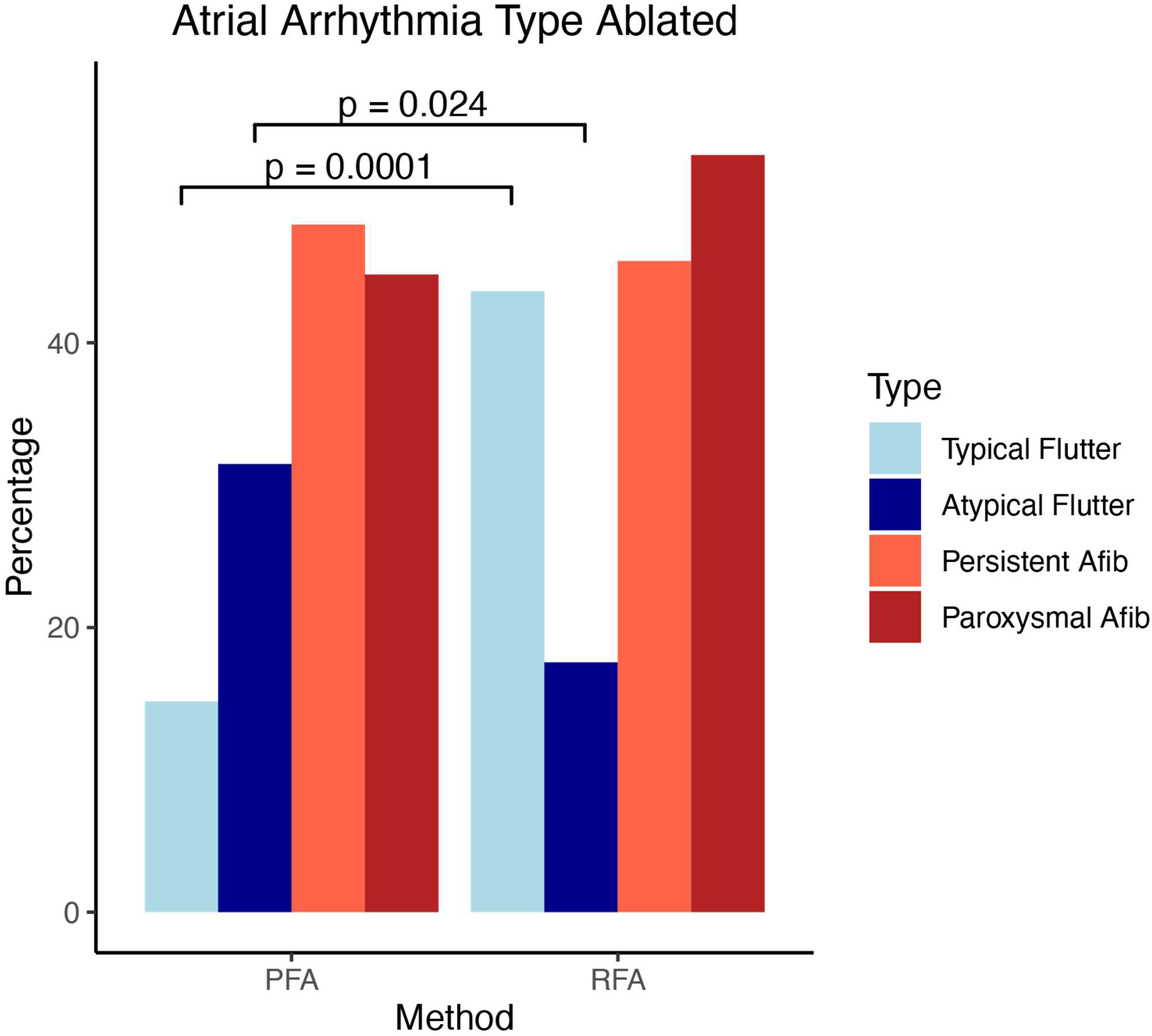
Atrial arrhythmia type ablated. *P* values were calculated using the Pearson’s χ^2^ test. PFA = pulsed-field ablation, RFA = radiofrequency ablation

### Recurrent Atrial Arrhythmias Following Repeat Ablation

Among PFA-treated patients, 28% had a recurrence of AF or AFL, compared to 55% of RFA-treated patients. Median follow-up was 9.3 (IQR = 5.8) months. Time to recurrence of PFA-treated patients was significantly longer than RFA patients (p=0.03) by Kaplan-Meier analysis (Fig 2A). When examining recurrence rates within the blanking period, there were greater recurrences among the RFA group (16.6% PFA vs. 29.0%, RFA p=0.3). Among those with early recurrence, 64% had AF/AFL recurrences in longer-term follow-up). After controlling for comorbidities (history of stroke, CHF, age, total CHADSVASC score, persistent vs paroxysmal AF, and sex (Figure 3)), only use of PFA for repeat ablation remained significantly associated with decreased AF/AFL recurrence (HR 0.4, 95% CI 0.2-0.9), and only presense of persistent AF (HR 2.27, 95% CI 1.19-4.33) was associated with increased risk of AF/AFL recurrence. Multivariate analysis was conducted in two segments. The first analysis included only CHADSVASC score and type of AF. Here, PFA demonstrated lower recurrence (HR 0.4, 95% CI 0.2-0.9, p = 0.04). This was further reinforced with the addition of all other demographic and risk factors considered (HR 0.4, 95% CI 0.2-0.9, p = 0.04).

**Figure 2A:**
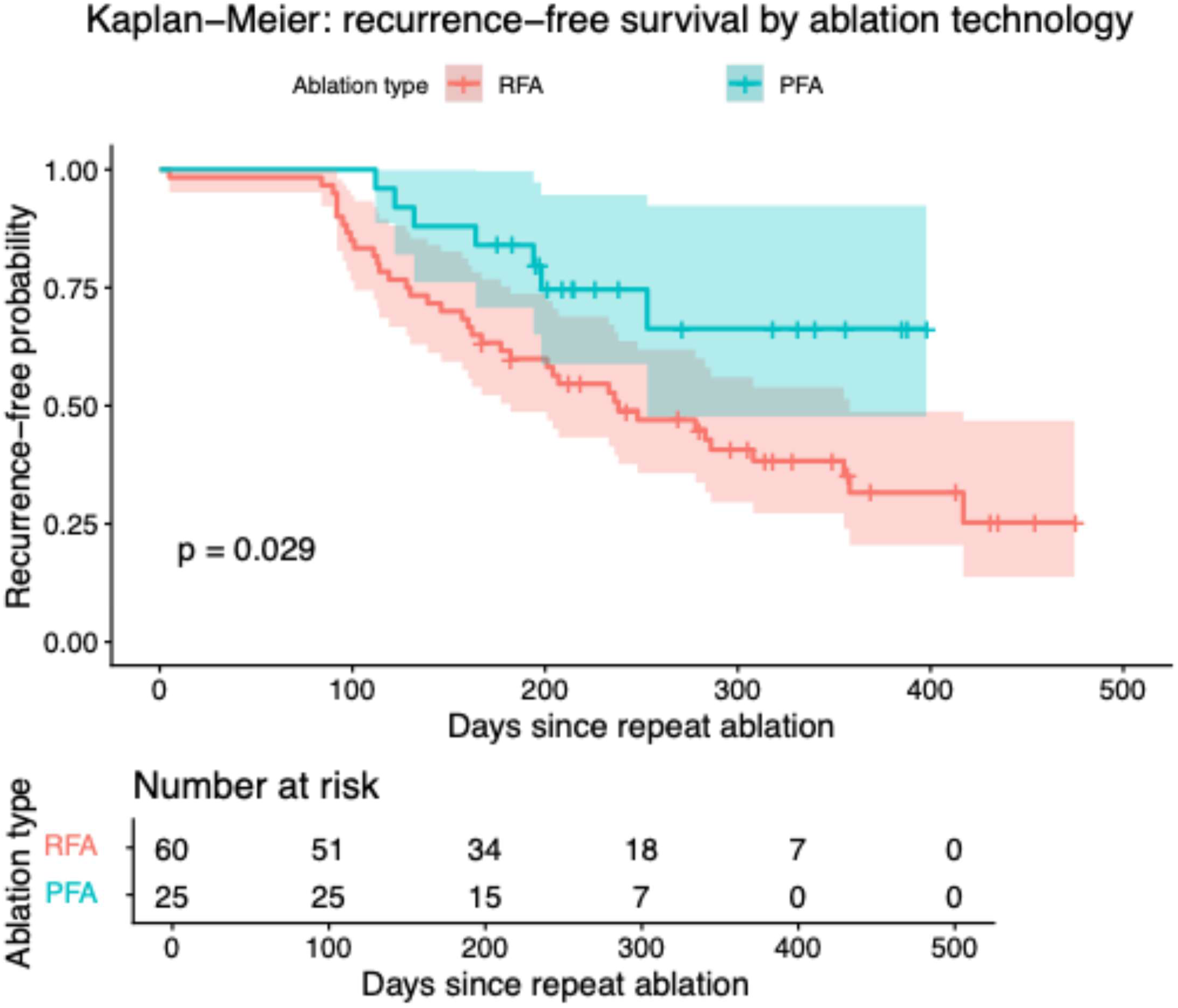
Recurrence of atrial fibrillation following redo-procedure with radiofrequency versus pulsed field ablation. Kaplan-Meyer curves comparing the probability of recurrence-free survival between PFA (n=54) versus RFA (n=62).

**Figure 2B:**
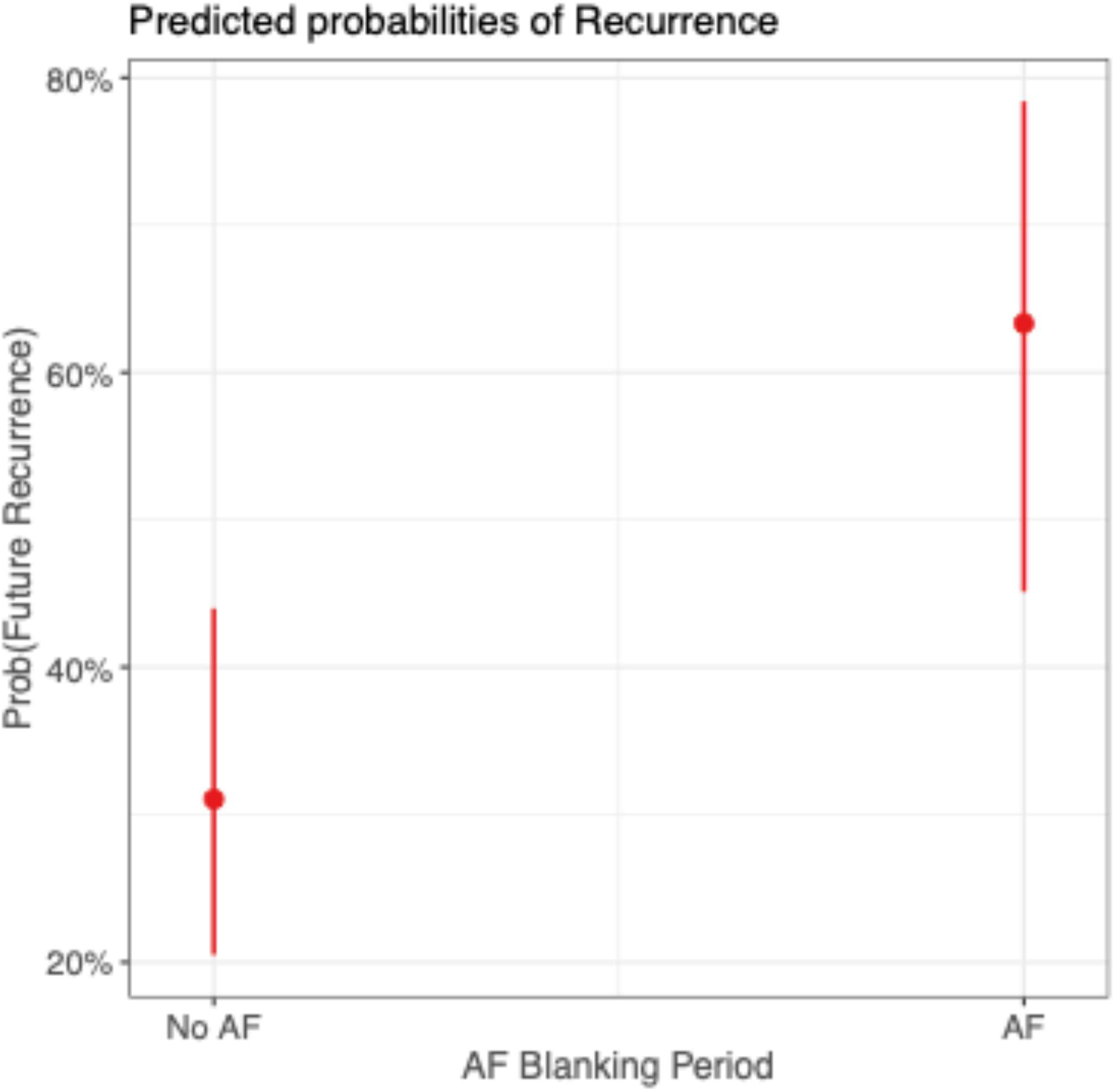
Probability of recurrence given atrial fibrillation within blanking period (90-days) Model of recurrence given AF within 90-day blanking period. Calculated using logistic binomial regression for future recurrence across both PFA and RFA. AF = atrial fibrillation

**Figure 3:**
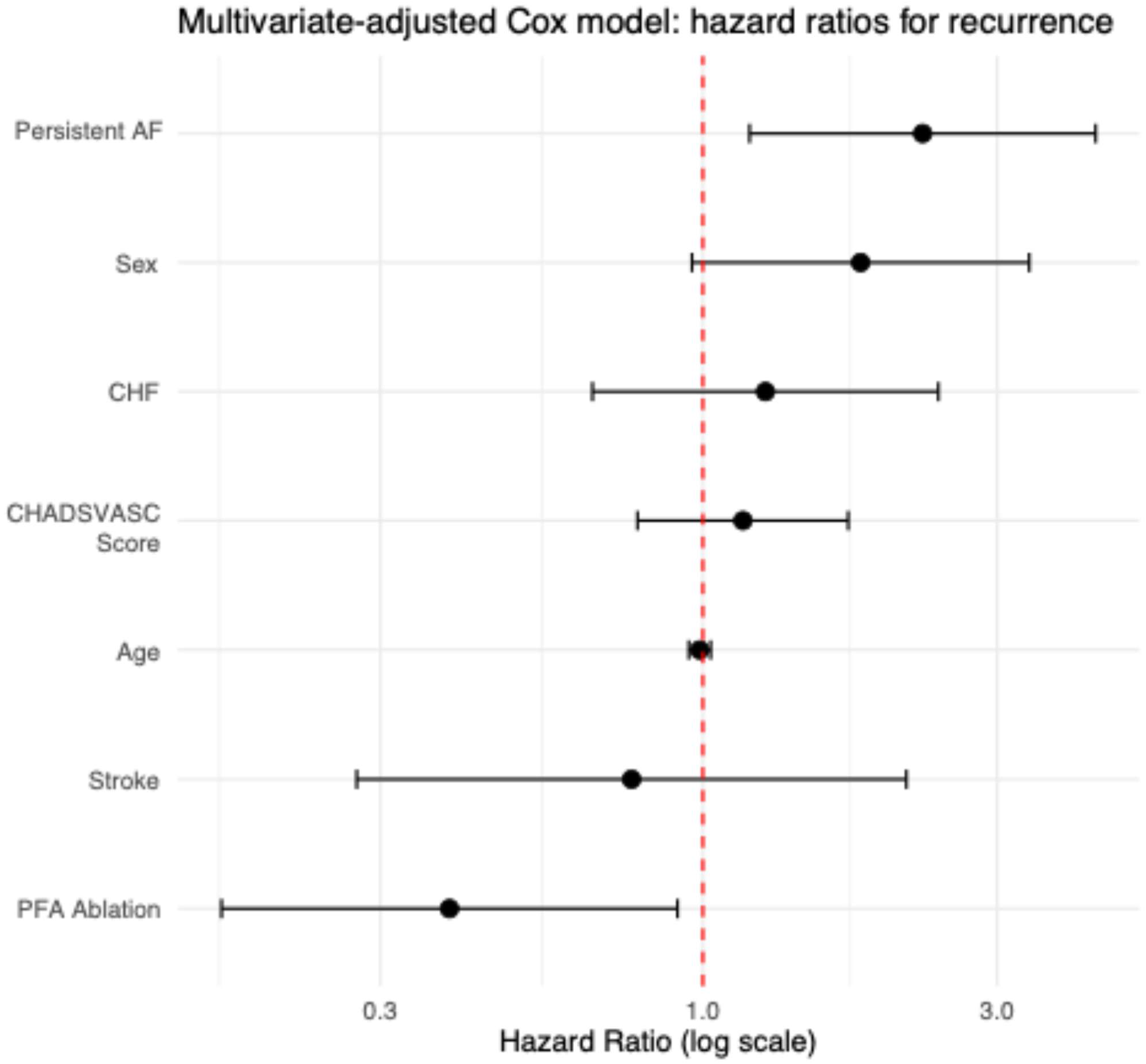
Multivariate Cox analysis and hazard ratios for risk-factors related to recurrence. Multivariate analysis to identify individual risk factors contributing to recurrence of arrhythmia following ablation. Hazard ratios <1 favor lower chance of recurrence of atrial fibrillation. Error bars represent 95% confidence intervals.

### Adverse Events

Adverse event rates were similar between PFA and RFA procedures and were predominated by vascular access complications (Table 3). Of note, one patient experienced sinus node dysfunction following SVC isolation with PFA and required pacemaker implantation. Similar proportions of patients experienced stable pericardial effusions which were managed conservatively, while one required pericardiocentesis for pericardial tamponade in a procedure utilizing RFA.

**Table 3:**
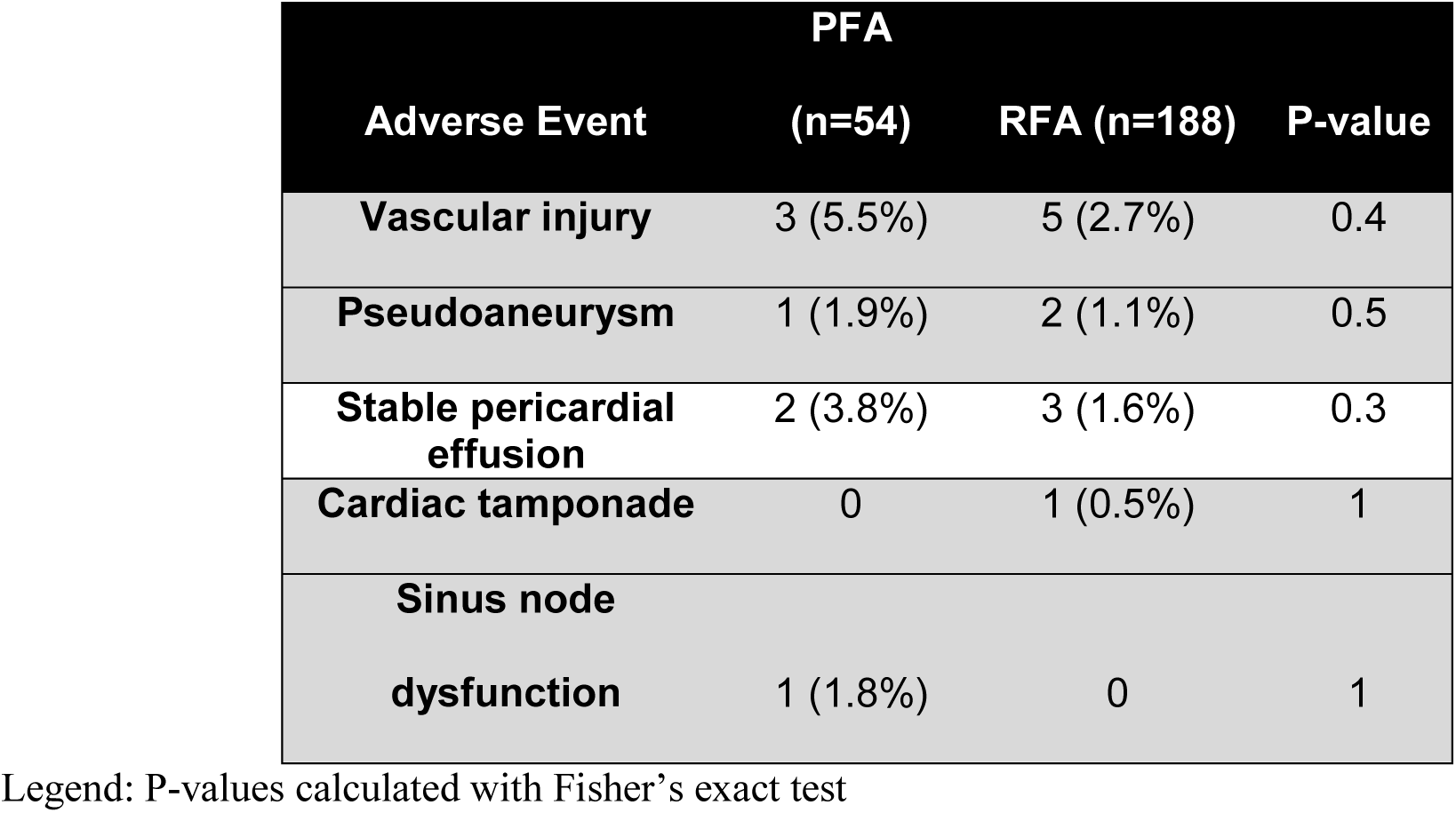
Adverse events associated with ablation.

## Discussion

This study is the largest to-date to compare the efficacy and outcomes for patients, in whom the index AF procedure was performed with RFA, undergoing repeat AF ablation with either PFA or RFA. Patients in whom PFA was chosen for repeat AF ablation were younger and with fewer baseline comorbidities, resulting in lower CHA2DS2-VASc scores, although had larger LA sizes and more often had atypical AFL along with recurrent AF. PFA procedures were shorter than RFA procedures, although fluoroscopy utilization was a mean of 5 min longer. In the context of these differences, the outcomes of AF/AFL-free survival was longer in the PFA compared to RFA group, although recurrences in the traditionally accepted 90-day blanking period following ablation were similar between groups. When incorporating health risk factors such as CHF, stroke, age, etc. PFA was shown to have a lower chance of recurrence compared with RFA.

Complication rates, which were predominated by vascular access events and not specific to energy modality, were similar across both PFA and RFA groups, as well as in other experiences reported.^6,7^ The notable exception is the patient who developed pacemaker-requiring sinus node dysfunction in the process of SVC isolation with PFA. This example speaks to an inherent limitation of initially clinically available PFA platforms, which were designed for single-shot PV isolation, as well as the need for operators to be wary of the unknown safety of distance that PFA can be applied without adversely affecting structures such as the sinus node, atrioventricular node, or healthy tissue.

Clinical outcomes from this study differed from previous data analyzing repeat catheter ablation with PFA vs RFA (n=185).^21^ Isenegger et al. found similar recurrence free survival in both PFA and RFA groups. On 1-year follow-up, recurrence-free survival of AF was observed to be 70% for PFA and 80% for RFA. 1-year Kaplan-Meier estimates demonstrated recurrence-free survival rate of 61% PFA vs 69% for the RFA group. Our study demonstrated lower recurrent AF/AFL following repeat PFA ablation (28% vs. 55%). Additionally, Kaplan-Meier analysis after a 90-day blanking period demonstrated a longer recurrence-free survival among PFA-treated patients. Of note, procedure characteristics and safety were similar across both studies, with minimal adverse events overall and PFA requiring increased fluoroscopy utilization. Differences between studies may be due to our study utilizing PFA across a broader region rather than pulmonary vein isolation alone. Patients in this study received a greater number of non-PVI ablation lesions to treat reccurent AF compared to previous use of PFA, which mainly utilized PWI as an additional lesion set.^21^

Badertscher et al. evaluated the value of PWI in addition to PVI for repeat catheter ablation of AF.^22^ In that propensity-matched study, 122 patients undergoing redo PVI alone were compared with 122 patients undergoing redo PVI plus PFA-based posterior wall isolation. The authors found no significant difference in arrhythmia-free survival at a median follow-up of 370 days. Results were comparable between groups with 69% in PVI-only and 72% in PVI + PWI group. Median procedure duration for PVI only groups was significantly shorter compared to the PVI+ group as well as PFA procedures from our study. However, outcomes observed in our study were not necessarily due to routine posterior wall isolation. In our study, roof/posterior LA ablation was uncommon and similar between both PFA and RFA groups. In contrast, additional lesions sets typically targeted other atypical atrial flutter regions, particularly mitral annular or CTI-dependent flutter. Together, these results support that non-PV ablation with PFA may be associated with more favorable outcomes in selected redo-ablation patients.

To summarize, these findings demonstrate that PFA provides advantages compared to RFA in the context of recurrent AF ablation. Of note, recurrent PFA ablations often included additional lesion sets aside from PVI, potentially contributing to lower recurrence rates. Previous studies have also demonstrated PFA to be a more efficient method of ablation and have noted PFA to have more durable PVI lesions. Taken together, these studies support PFA as a safe and efficient tool for repeat ablation, but they also suggest that improved outcomes may depend on appropriate patient selection and arrhythmia-specific targeting rather than empiric expansion of lesion sets.

Despite these promising findings, several important questions remain regarding the optimal approach to repeat AF ablation. While our data suggest that additional lesion sets may contribute to improved outcomes in PFA patients, the relative durability of non-PVI lesions created by PFA has not been thoroughly characterized. Current evidence on lesion strength is largely confined to PVI, and it remains unclear whether this translates equivalently to posterior wall isolation, mitral isthmus lines, cavotricuspid isthmus ablation, or other substrate-based targets. Additionally, as use of PFA is expanding, questions regarding safety and efficacy continue to arise. Future work will need to be done over a longer follow-up period to characterize these findings.

## Limitations

This study has several limitations to acknowledge. It was a nonrandomized, retrospective single center study, with associated limitations, including selection bias and unmeasured confounding, and the relatively small cohort may have limited ability to detect statistically significant differences. However, there was little difference in baseline characteristics of PFA and RFA groups. Additionally, as recurrent AF/AFL was not assess uniformly using continuous monitoring, rates of AF/AFL recurrence could have been underestimated; we would have expected for this bias to be similar in both PFA and RFA groups, however. Further this study represents our early institutional experience with pulsed field ablation using a single catheter platform, with proprietary PFA waveform algorithms. Therefore, these findings can not be generalized to other PFA systems, catheter designs, or more contemporary ablation workflows. In addition, non-pulmonary vein ablation performed with this catheter was off-label, which should be considered when interpreting the safety, feasibility, and efficacy of these procedures.

## Data Availability

Data analyzed in this manuscript, other than already included, can be made available upon reasonable request.

## Summary

Despite limitations, our study is unique in identifying the potential advantage of PFA over RFA for repeat ablation of recurrent AF/AFL.

## FINANCIAL MATERIAL & SUPPORT

No funding or other support was required for this study.

## CONFLICTS OF INTEREST TO DECLARE

Dr. Tzou has received research funding from or served as consultant/advisor for American Heart Association, American College of Cardiology, Abbott, Bayer, Johnson & Johnson Med Tech, Boston Scientific, Field Medical, Medtronic, Physcade, Thermedical, and Varian.

Dr. Sabzwari has received speaker’s honorarium from Abbott, and consultation for Medtronic.

Dr. Garg has received speaker honoraria or research funding from Abbot, & Johnson MedTech, and Bayer

Dr. Tumolo has recieved speaker honoraria with AcademicCME and educational programming from Biosense Webster, Boston Scientific, and Abbott.

Dr. Zipse has received consultant positions from Philips, Medtronic, and Boston Scientific. He has also been assisted with educational programming with Johnson & Johnson MedTech

All other authors have reported that they have no relationships relevant to the contents of this paper to disclose.

## Nonstandard Abbreviations and Acronyms

PFA: pulsed field ablation
AF: atrial fibrillation
RFA: radiofrequency ablation
PWI: posterior wall isolation
AAs: atrial arrhythmias
CA: catheter ablation
PVI: pulmonary vein isolation
AFl: atrial flutter
AT: atrial tachycardia
LA: left atrium
RA: right atrium
SVC: superior vena cava
IVC: inferior vena cava

